# Performance comparison of reusable versus disposable colonoscopes:a non-inferiority Trial

**DOI:** 10.1101/2022.11.20.22282561

**Authors:** Mingtong Wei, Chenghai Liang, Huaqiang Ruan, Guolin Liao, Peng Peng, Xin Li, Jun Zou, Shiquan Liu, Ge Cao, Mengbin Qin, Jiean Huang

## Abstract

**Objective:** We herein compared the performance of reusable and disposable colonoscopes in patients scheduled to undergo colonoscopy with a view of preventing patient cross-infection, protecting the safety of clinical medical staff, reducing the risk of infection, and minimizing the decontamination process, particularly during the coronavirus disease 2019 pandemic.

**Methods:** We randomly divided patients meeting the enrollment criteria into reusable and disposable colonoscopy groups; the success rate of photographing customary anatomical sites with a non-inferiority margin of -8% was the primary endpoint. Secondary endpoints were the adenoma detection rate, operation time, endoscopic image quality score, endoscopic mucosal resection (EMR) success rate, and adverse events.

**Results:** We recruited patients who were treated using reusable or disposable (n = 45, each) colonoscopes. Both groups had 100% success rate for capturing images of customary anatomical sites, with no between-group differences. The lower limit of 95% CI was - 7.8654%, which was greater than the non-inferiority threshold of -8%. The disposable group had a significantly lower average image quality score (26.09 ±1.33 vs. 27.44±0.59, *P* < 0.001) than the reusable group. The groups did not significantly differ in maneuverability, safety, or device failure/defect rate. The *en-bloc* EMR success rate was 100% in both groups. EMR took significantly longer in the disposable group (466.18 s±180.56 s *vs*. 206.32 s±109.54 s, *P* < 0.001). The incidence of EMR-related bleeding and perforation did not significantly differ between the groups.

**Conclusions:** Disposable colonoscope endoscopy is safe and feasible for endoscopy examinations and EMR.

## Introduction

With the introduction of new instruments, procedures, and accessories in medicine, the field of gastrointestinal (GI) endoscopy has significantly expanded, and in Japan and the United States alone, more than 15 million GI endoscopies are performed every year [1,2]. Notably, GI endoscopies have wide implementations in the diagnosis, treatment, and postoperative follow-up of digestive tract diseases. They can be used to directly visualize the affected part of the GI and diagnose the disease and can be combined with several surgical tools for treatment. At present, the same GI endoscopes are used for consecutive patients, because of which they need to be cleaned, disinfected, and sterilized after every use. The cleaning and disinfection process is cumbersome and warrants a dedicated cleaning and disinfection site and specialized cleaning and disinfection equipment [3]. It is not possible to completely prevent endoscope reuse-related infections despite detailed and strict guidelines for cleaning and disinfecting endoscopes [4,5,6]. In this study, we evaluate the safety, feasibility, and performance of disposable colonoscope endoscopy with a focus on preventing cross-infection among patients and ensuring the safety of clinical medical staff.

## Materials and methods

### Disposable endoscopy technique

The disposable colonoscope was provided by HuiZhou Xzing Technology Co. Ltd. Disposable endoscopy (XZING-C200B) parameters were as follows: field of view, 110°; direct field observation; depth of field, 3–100 mm; working length, 1300 mm; operating channel inner diameter, ≥φ3.0 mm; and angle ranges: up and down, ≥180º and left and right, ≥160°. Three gastroenterologists with a collective experience of >10,000 cases of endoscopy and treatment holding positions of deputy chief physician or above were trained for and mastered the operating methods of medical disposable colonoscope endoscopy systems. The reusable colonoscopes used were OLYMPUS CF-HQ290ZI, CF-H290I, and PCF-TYPE-Q260AZI.

### Patient selection and defined variables

#### 1. Patient selection

We performed a trial which was approved by the Ethics Committee of The Second Affiliated Hospital of Guangxi Medical University on March 23, 2021 (No. KY-0108). Ninety patients were recruited into the group. Trial Registration: The trial is registered at the Chinese Clinical Trial Registry (http://www.chictr.org.cn) and Registration number: ChiCTR2100045084. These patients were 18-75-year-old men and women who underwent colonoscopy under anesthesia or sedation and voluntarily participated in this trial with signed written informed consent. We excluded patients who were contraindicated to undergo colonoscopy and patients who had 1) a thoracic–abdominal aortic aneurysm, 2) high-grade spinal malformations, 3) severe cardiovascular and cerebrovascular diseases, 4) acute radiation colitis, 5) a huge diverticulum in the digestive tract, 6) an advanced tumor with abdominal metastasis and/or obvious ascites, 7) peritonitis or gastrointestinal perforation; 8) an existing systemic bleeding disease or abnormal coagulation function and bleeding, and 9) severe intellectual disability or mental illnesses keeping them from cooperating during the procedure. Furthermore, we excluded pregnant and lactating women, patients undergoing emergency gastrointestinal endoscopy and/or treatment, patients with a history of colonic surgery, patients who were participants in other clinical trials 1 month before screening, patients with a history of allergy to anesthetics, and patients considered unsuitable for participation by the investigator.

#### 2. Main evaluation index

##### 2.1 Primary measure

###### Evaluation method

The entire procedure was recorded, and colonoscopy images for the following anatomical locations were observed: the lower rectum 2 cm above the anal verge, the middle part of the sigmoid colon, the descending colon of the splenic flexure, the transverse colon posterior to the splenic flexure, the transverse colon anterior to the hepatic flexure, the ascending colon posterior to the hepatic flexure, and the ileocecal region. Single or multiple representative images were acquired for each site. Furthermore, the colonoscopy report contained photographs and records of all abnormalities identified. For the evaluation, we used the clearest image of each of these seven sites. The image evaluation adopted a two-person (attending physicians) back-to-back evaluation method. In case of inconsistent results, a third investigator was invited, and agreement of two of the three investigators was considered for final evaluation.

###### Evaluation criterion

The image quality was considered acceptable only if at least one representative image of the seven anatomical markers had no missing sites with a clear image. Under any other condition, the image quality was considered unacceptable. The equation below shows how the percentage of acceptable image quality cases was quantified.

Acceptable image quality (%) = number of subjects in each group with acceptable image quality (n) ÷ number of subjects in each group × 100%.

##### 2.2 Secondary measures

###### 2.2.1 Colonoscopy image quality score

Image quality was rated on a scale of 0–4 on the basis of their completeness and clarity: 0, missing sites/unclear images; 1, no missing sites but slightly unclear images; 2, no missing sites and relatively clear images; 3, no missing sites and clear images; and 4, no missing sites and extremely clear images [7]. For each subject, the total score of the above images was calculated.

###### 2.2.2 Cecal intubation rates

Cecal intubation was defined as advancing the colonoscope tip close to the ileocecal valve to allow visual access to the entire cecal caput, including the medial wall of the cecum between the appendiceal orifice and the ileocecal valve [8]. The equation below shows how the cecal intubation rate was calculated.

Cecal intubation rate (%) = number of subjects with cecal intubation in each group (n) ÷ number of subjects in each group × 100%.

###### 2.2.3 Operative time of colonoscopy

The withdrawal time, indicative of the time taken to retract the colonoscope from the ileocecal region until complete withdrawal [excluding the time taken for additional steps during the procedure, such as pathological biopsy/endoscopic mucosal resection (EMR)], was recorded.

###### 2.2.4 Detection rates for adenomas, polyps, and diminutive polyps

The adenoma detection rate (ADR) is the percentage of patients detected as having one or more adenomas among the patients who underwent complete screening via colonoscopy. Polyp detection rate (PDR) represents the number of patients with ≥1 polyp removed during screening colonoscopy [9]. In addition, diminutive polyps (<5 mm in diameter) are typically the vast majority of polyps found during screening colonoscopy.

###### 2.2.5.1 Procedure time of EMR

The EMR technique involves locally injecting dilute methylene blue (dilution concentration 5‰) into the submucosal layer. This is followed by snaring and resection, and clipping is used to prevent delayed bleeding after EMR. The EMR procedure begins with submucosal injection and ends with complete resection of the lesion. The size, morphology, site, and access (SMSA) scores and levels of polyps were recorded [10].

###### 2.2.5.2 En-bloc resection rate

Successful en-bloc resection of polyps was judged as effective. Incomplete tumor resection or the presence of residual tumor tissue was judged as invalid.

En-bloc resection rate (%) = number of subjects with successful en-bloc resection in each group (n) ÷ number of subjects in each group × 100%.

###### 2.2.5.3 Adverse events of EMR

Perforation: Perforation was indicated by severe abdominal pain, peritoneal irritation, and the presence of freeing gas under the diaphragm identified on X-ray or abdominal CT.

Immediate bleeding: A non-self-limiting bleeding lasting >60 s was defined as acute intraprocedural bleeding [11].

Delayed bleeding: Delayed bleeding was defined as a drop in hemoglobin of ≥2 g/dL and bright red rectal blood loss [any postprocedural blood loss from the anus warranting re-hospitalization, emergency room consultation, prolonged hospital stay, blood transfusion, or re-intervention (repeat endoscopy, angiography, or surgery)] [11]. Patients who underwent EMR were followed up for 1 week, 2 weeks, and 1 month to evaluate whether there was delayed bleeding.

###### 2.2.6 Acceptable clinical operability

The operator-evaluated operability was assessed on the basis of knob operation, acute angle adaptability, lesion biopsy, aspiration operation, water delivery operation, and suction function.

Evaluation criteria: Each item was rated as A (high), B (fair), or C (low). Acceptable clinical operability was indicated by both image quality and operability being rated A or B. If this condition was not met, clinical operability was considered unacceptable.

###### 2.2.7 Equipment failure/defects rate

The occurrence of device failure, such as water jet malfunction and image interruption, during the procedure was recorded.

## Statistical analysis

We used SAS 9.4 for statistical analyses. Measurement data were analyzed using the signed rank-sum test or paired t-test for intragroup comparisons and the Wilcoxon rank-sum test or t-test for intergroup comparisons; count data were analyzed using the chi-squared test or Fisher’s exact test for intergroup comparisons.

## Results

### 1. General information

90 patients were recruited into the group and completed this clinical trial. The parameters of sex and age did not significantly differ between the groups (Table 1).

**Table 1.** Demographic and clinical data of patients

|  | Disposable | Reusable | *P* |
| --- | --- | --- | --- |
| Age (years, mean, range) | 43.16 ± 14.41 | 43.49 ± 13.10 | 0.909 |
| Sex, n |  |  | 1.000 |
| Male | 24 | 24 |  |
| Female | 21 | 21 |  |
| History of allergy to anesthetics, n |  |  | 1.000 |
| Yes | 0 | 0 |  |
| No | 45 | 45 |  |
| History of gastrointestinal surgery, n |  |  | 1.000 |
| Yes | 0 | 0 |  |
| No | 45 | 45 |  |
| Abdominalgia, n |  |  | 1.000 |
| Yes | 14 | 15 |  |
| No | 31 | 30 |  |
| Bloating, n |  |  | 0.367 |
| Yes | 17 | 12 |  |
| No | 28 | 33 |  |
| Diarrhea, n |  |  | 1.000 |
| Yes | 4 | 5 |  |
| No | 41 | 40 |  |
| Prior history of polyp, n |  |  | 0.399 |
| Yes | 20 | 25 |  |
| No | 33 | 29 |  |
| Constipation, n |  |  | 0.606 |
| Yes | 8 | 11 |  |
| No | 37 | 34 |  |
| Bowel preparation score | 7.78 ± 1.30 | 7.49 ± 0.99 | 0.238 |

### 2. Effectiveness evaluation

#### 2.1 Primary measure: acceptable image quality

In both groups, the image quality was rated as acceptable for all patients [45/45, 95% confidence interval (CI): 0.9213, 1.0000] when judged on the basis of the primary endpoint with a non-inferiority margin of -8%. There were no between-group differences, and the lower limit of the 95% CI was -7.8654% (-7.8654%–7.8654%), which was greater than the non-inferiority threshold of -8%, indicating that image quality in the disposable group was not inferior to that in the reusable group (Table 2).

**Table 2.**
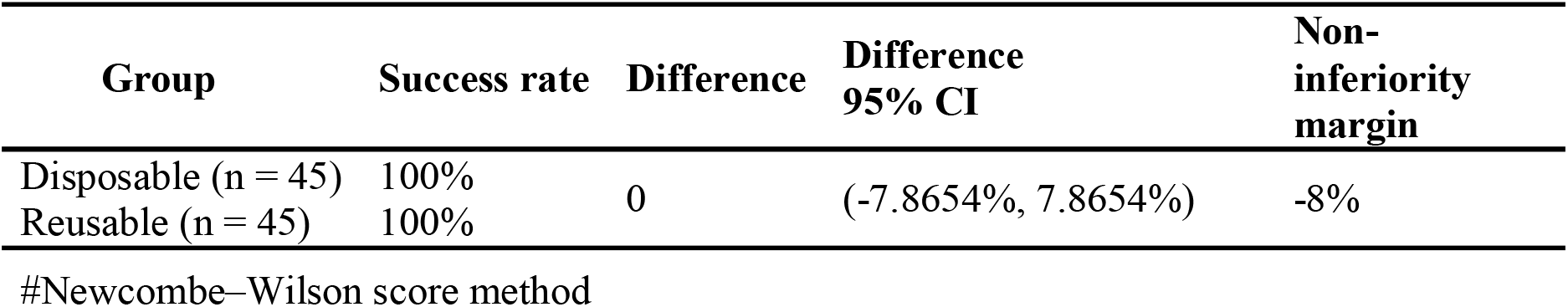
The success rate of photographing iconic anatomical sites in the two groups

#### 2.2 Secondary measures

##### 2.2.1 Colonoscopy image quality score

The mean image quality scores in the disposable and reusable groups significantly differed and were 26.09 ± 1.33 and 27.44 ± 0.59, respectively (*P* < 0.001). For site scores, in both groups, all sites were rated as 3 or above; however, site scores were lower in the disposable group than in the reusable group for the middle part of the sigmoid colon, the transverse colon posterior to the splenic flexure, the transverse colon anterior to the hepatic flexure, and the ileocecal region, and the differences were statistically significant (*P* < 0.05) (Table 3).

**Table 3.**
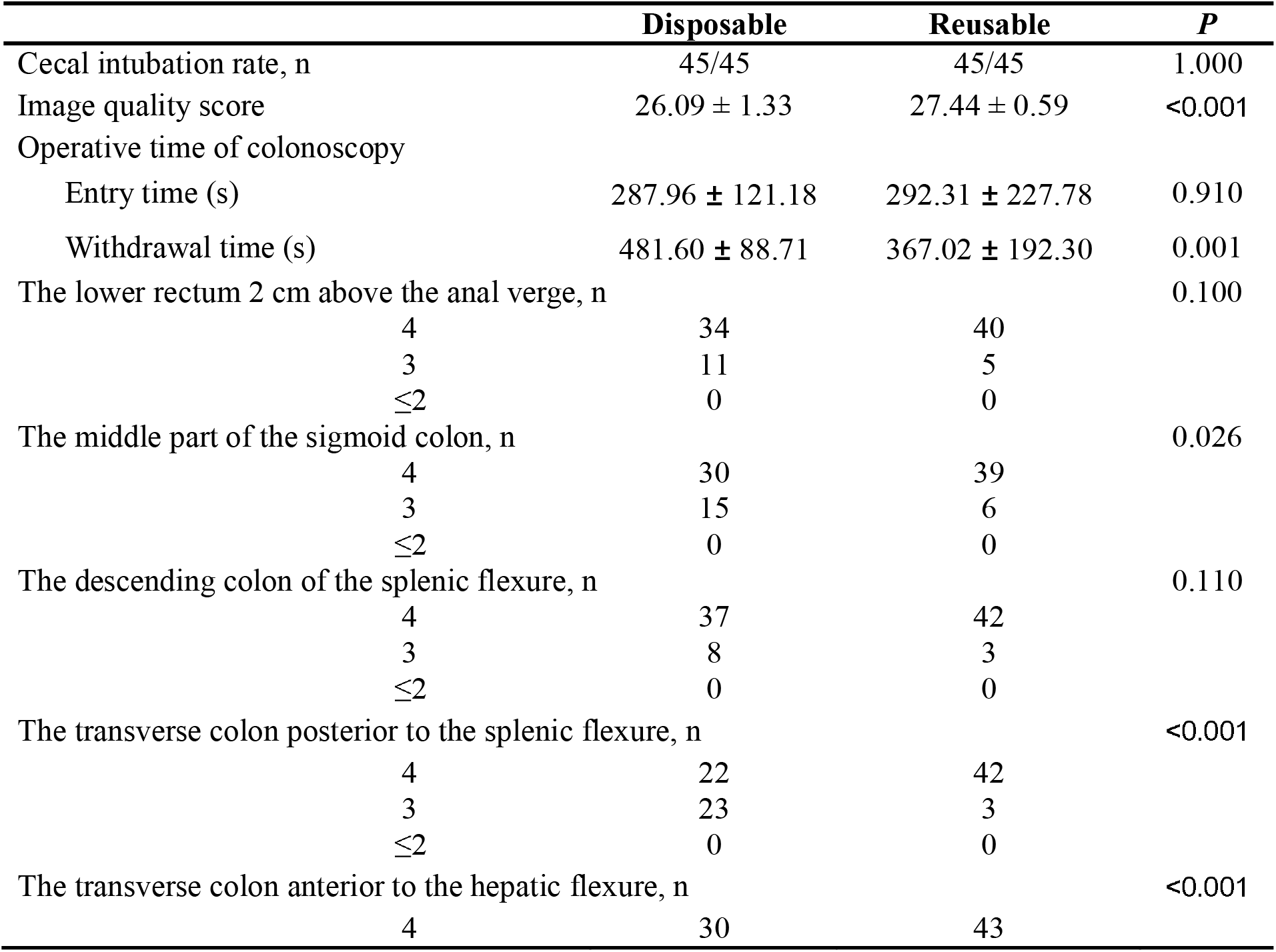
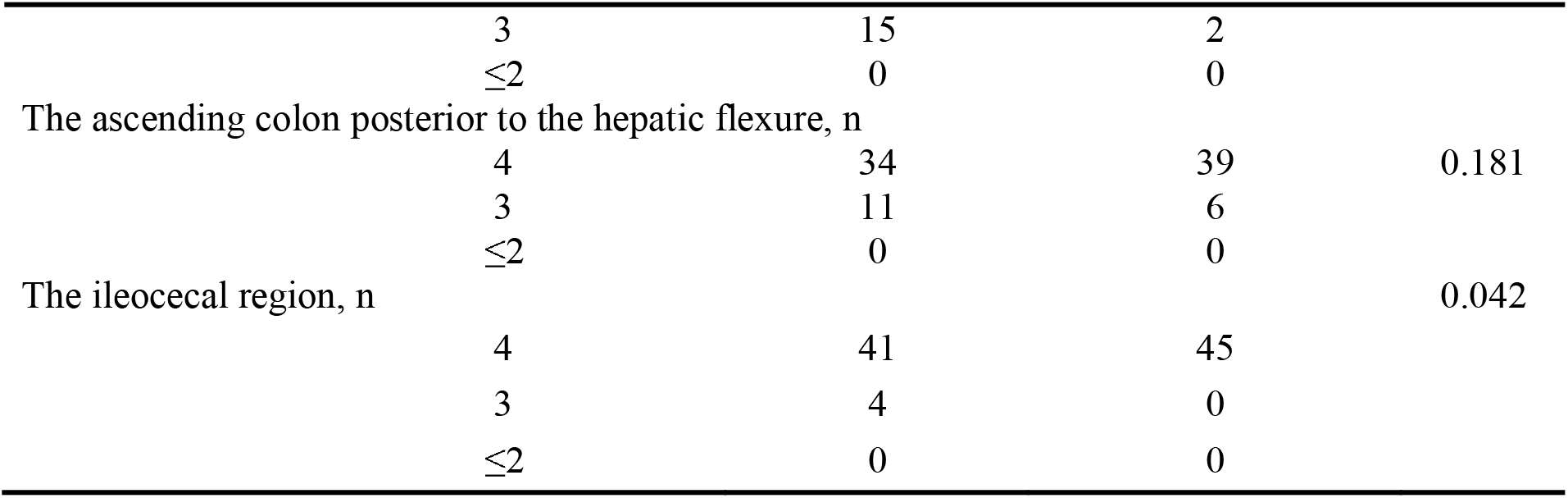
The secondary measures of both groups

##### 2.2.2 Cecal intubation rates

The cecal intubation rates were 100% (45/45) in both groups (Table 3).

##### 2.2.3 Adenoma detection rate

In the experimental group, 23 polyps were found in 21 patients (detection rate, 46.7%). In the control group, 25 polyps were found in 22 patients (detection rate, 48.9%); the ADR values were 14/45 (31.1%) and 11/45 (24.4%) in the experimental and control groups, respectively (Table 4).

**Table 4.** The efficacy and complications of EMR in both groups

|  | **Disposable** | **Reusable** | ***P*** |
| --- | --- | --- | --- |
| Polyp detection, n (%) | 21/45(46.7%) | 22/45(48.9%) | 1.000 |
| Total number of polyps, n | 29 | 31 | NS |
| Hyperplastic polyp, n | 15 | 20 |  |
| Tubular adenoma, n | 11 | 7 |  |
| Tubulovillous adenoma, n | 2 | 2 |  |
| SSL, n | 0 | 1 |  |
| TSA, n | 1 | 0 |  |
| Juvenile polyp, n | 0 | 1 |  |
| Adenoma detection, n | 14/45 | 11/45 | 0.638 |
| Diminutive polyp detection, n (%) | 11/45 (24.4%) | 23/45 (51.1%) | 0.016 |
| Procedure time of EMR(s) | 466.18 ± 180.56 | 206.32 ± 109.54 | 0.000 |
| En-bloc resection, n | 19/19 | 17/17 | 1.000 |
| SMSA score of colonic polyps | 5.00 ± 0.69 | 4.89 ± 0.93 | 0.689 |

| AEs |  |  |  |
| --- | --- | --- | --- |
| Immediate bleeding, n | 1/19 | 1/17 | 1.000 |
| Delayed bleeding, n | 1/19 | 1/17 | 1.000 |
| Perforation, n | 0/19 | 0/19 | 1.000 |

SSL: sessile serrated lesion; TSA: traditional serrated adenoma; AEs: adverse events; SMSA score: the size, morphology, site, and access score

##### 2.2.4 Efficacy and complications of EMR

EMR was performed for 19 polyps in the experimental group and 17 polyps in the control group; the overall en-bloc resection rate was 100%. The between-group difference in SMSA scores was not statistically significant; however, the procedure of EMR lasted significantly longer in the experimental group than in the control group (466.18 ± 180.56; 206.32 ± 109.54; *P* < 0.05).

Both groups had one case each of sudden bleeding during EMR, and the bleeding stopped after thermal coagulation or hemostasis with titanium clips; one case in the experimental group occurred DB in 48 h after EMR, and one case in the control occurred DB in 24 h after EMR. None of the subjects were identified to have perforations at 24 h after the procedure (Table 4).

##### 2.2.5 Acceptable clinical operability

The operation evaluation (rating: A or B) was 100% in both groups for the following: auxiliary features (water supply, air supply, and suction) and flexibility (knob operation, body rigidity, and sharp angle adaptability). For the rating of operability (A, B, or C), no significant differences were observed between the groups in terms of air supply (Table 5).

**Table 5.** The operation evaluation in the two groups

|  | Disposable | Reusable | *P* |
| --- | --- | --- | --- |
| **Flexibility** |  |  |  |
| **Body rigidity** |  |  | 0.550 |
| A: Moderate rigidity, good operability | 37 | 40 |  |
| B: Slightly Soft or Slightly hard, general operation | 8 | 5 |  |
| **Knob operation** |  |  | 0.242 |
| A: Flexible | 42 | 45 |  |
| B: Fair, with certain resistance | 3 | 0 |  |
| **Sharp angle adaptability** |  |  | 0.056 |
| A: Good: the tip of the scope can easily pass | 40 | 45 |  |
| B: Fair: the tip of the scope can pass but with relatively less ease | 5 | 0 |  |
| **Auxiliary features** |  |  |  |
| **Air supply** |  |  | 0.494 |
| A: Operation is responsive immediately and supplies moderate air supply. | 43 | 45 |  |
| B: Operation is responsive but not immediate with more or less air supply. | 2 | 0 |  |
| **Water supply** |  |  | 1.000 |
| A: Operation is responsive immediately and the scope can be effectively cleaned. | 44 | 45 |  |
| B: Operation is responsive but not immediate and the scope can be effectively cleaned. | 1 | 0 |  |
| C: Operation is not responsive or with no water supply. | 0 | 0 |  |
| **Suction** |  |  | 1.000 |
| A: Operation is responsive, and the fluid level drops or the bowel retracts immediately. | 44 | 45 |  |
| B: Operation is responsive, and fluid level drops or the bowel retracts but not immediately. | 1 | 0 |  |
| C: Operation is not responsive with no fluid level drops or bowel retraction. | 0 | 0 |  |

##### 2.2.6 Device failure/malfunction rate

In both groups, none of the devices showed failure/malfunction (rate: 0.0%).

##### 2.2.7 Safety evaluation

In both groups, none of the subjects in the study presented with hypotension, aspiration, or any other adverse event. This suggests that colonoscopy has a good safety profile causes no immediate or delayed harm to the patient.

## Discussion

Digestive endoscopy is a widely used procedure in clinical practice, and >10 million gastrointestinal endoscopy procedures are performed globally every year. Taking into account the 169 endoscopy units in China as of 2013, Zhang et al. [12] reported that the average number of gastrointestinal endoscopes per unit in 2013 was 9.3. There are typically eight reprocessing steps before reuse of a colonoscopy endoscope: precleaning, leak testing, manual cleaning, rinsing after cleaning, visual inspection for contamination, high-level disinfection, rinsing after high-level disinfection, and drying [13]. However, even after reprocessing and drying, gastrointestinal endoscopes were found to still have microbial growth, residual droplets, and biofilm formation [14], and therefore, completely preventing infections after endoscopy seems impossible despite the medical practices being guided by such rigorous guidelines [5, 15, 16]. Therefore, the patient-to-patient infection transmission risk should be eliminated by the design of disposable colonoscopes [17]. To our knowledge, this study is the first to use disposable colonoscopes designed by Huizhou Xzing Technology Co. Ltd. in a clinical controlled trial, However, this equipment has been used in animal experimental research and clinical research case reports in the past [18,19]. In the present study, the effectiveness, efficacy, and safety of these disposable colonoscopes were evaluated.

The quality of images acquired using these colonoscopes was good, and the images were complete. The image quality satisfied the clinical requirements, particularly with regard to the acceptable image quality rate. However, the image quality score was lower in the disposable group than in the reusable group, particularly in the middle part of the sigmoid colon, the transverse colon posterior to the splenic flexure, and the transverse colon anterior to the hepatic flexure; therefore, the performance of the disposable colonoscope does not appear to be as good as that of the reusable gastroscope, and we believe image clarity is the main differentiating factor in this regard. However, improvements can be made to the image quality and maneuverability in the future.

The operative time of endoscopy is affected by the flexibility of the endoscope and the proficiency of the operator, and it is also influenced by the air/water supply and suction during the inspection process [18,20]. In our study, the disposable group had a longer operative time (either the endoscope withdrawal time or the time required for polyp EMR) than the reusable group, but the flexibility rate, as determined by body rigidity, knob operation, and sharp angle adaptability of the scope, was acceptable at 100% in both groups, indicating good operability and flexibility. Similarly, the auxiliary features, namely air supply, water supply, and suction, were also found to be acceptable at 100% in both groups, which is indicative of good air and water supply and suction. This allows for effective cleaning of the scope (self-cleaning) and removal of residual liquid or food from the site (site cleaning), thus ensuring successful and safe examination or treatment.

The ADR is the most closely related metric to the development of post-colonoscopy colorectal cancer or interval colorectal cancer and is an indicator of colonoscopy quality; another valuable indicator of colonoscopy quality is the polyp detection rate, which is much simpler to calculate than the ADR [9,21]. Approximately half of diminutive polyps (polyp size, <5 mm in diameter), which are the vast majority of polyps found during colonoscopy, are adenomatous and present with low risk of advanced neoplasia. In our study, the ADR was 14/45 (31.1%) and 11/45 (24.4%) in the disposable and reusable groups respectively, and the PDR was 46.7% and 48.9% respectively, with no significant between-group differences. This is indicative of quality colonoscopy examination or treatment. However, in our study, the detection rate of diminutive polyps was 11/45 (24.4%) and 23/45 (51.1%) in the experimental and control groups, respectively, and the difference was significant between the two group (*P* < 0.05). This finding suggests that diminutive polyp detection was slightly weaker using smart wavelength imaging (the disposable group) than using narrow band imaging (the reusable group).

Colonoscopy can help reduce the risk of colorectal cancer by facilitating the identification and removal of precancerous lesions (e.g., adenomas). According to the European Society of Gastrointestinal Endoscopy recommendations, the primary goal of EMR is to achieve a completely snare-resected lesion in the safest minimum number of pieces such that the margins are adequate and there is no need for adjunctive ablative techniques [22]. Our study showed successful en-bloc resection (100%) of colon polyps in both groups. We further found no statistically significant differences between the groups in terms of immediate bleeding, delayed bleeding, and perforation incidences, suggesting that disposable colonoscopes are as good as reusable colonoscopes.

There are some limitations to our study. First, this is a non-inferiority trial, and despite the image quality from disposable colonoscope being acceptable, the performance of the disposable colonoscope was not as good as that of the reusable colonoscope. However, improvements can be made to the image quality and maneuverability in the future. Furthermore, the polyp SMSA scores for EMR procedures were mainly distributed at SMSA levels 1–2 in our study, which could not be analyzed by subgroup discussion according to the SMSA grade because of the limited number of cases; this may be a direction to explore in the future.

In conclusion, disposable colonoscopy is effective and safe. It may be a favorable option under certain circumstances. It could serve as a personal protective equipment during the pandemic of COVID-19 or other infectious outbreaks.

## Data Availability

All data produced in the present work are contained in the manuscript

## Author’s contribution

Mingtong Wei, Huaqiang Ruan and Guolin Liao collected data, Chenghai Liang collected data, analyzed relevant information, and drafted the manuscript, Peng Peng, Xin Li, Jun Zou, Shiquan Liu, Mengbin Qin and Jiean Huang clinically managed the patient. Mengbin Qin and Jiean Huang designed the article, approved the final submission, and clinically managed the patient.

## Ethics declarations

### Declarations

Disclosures Mingtong Wei, Chenghai Liang, Huaqiang Ruan, Guolin Liao, Peng Peng, Xin Li, Jun Zou, Shiquan Liu, Ge Cao, Mengbin Qin, Jiean Huang have no conflicts of interest or financial ties to disclose.

